# Classification of Tauopathies from Human Brain Homogenates through Salt-Modulated Tau Amplification

**DOI:** 10.1101/2025.08.05.25331458

**Authors:** Alessia Santambrogio, Michael A. Metrick, Peifeng Xu, Shunsuke Koga, Bernardino Ghetti, Dennis W. Dickson, Byron Caughey, Michele Vendruscolo

**Affiliations:** Centre for Misfolding Diseases, Yusuf Hamied Department of Chemistry, University of Cambridge, Cambridge, UK; Laboratory of Neurological Infections and Immunity, Rocky Mountain Laboratories, Division of Intramural Research, National Institute for Allergy and Infectious Diseases, Hamilton, Montana, USA; Departments of Pathology and Laboratory Medicine, University of California at San Francisco, California, USA; Department of Neuroscience, Mayo Clinic, Jacksonville, Florida, USA; Department of Pathology and Laboratory Medicine, The University of Pennsylvania, Pennsylvania, USA; Department of Pathology, Indiana University School of Medicine, Indianapolis, Indiana, USA

## Abstract

Tauopathies are a heterogeneous group of neurodegenerative disorders characterised by the aggregation of the microtubule-associated protein tau in the brain. Recent advances in cryo-electron microscopy have revealed that tau aggregates adopt disease-specific structural conformations. However, translating these findings into practical diagnostic tools remains a challenge. We developed a salt-modulated real-time quaking-induced conversion (RT-QuIC) assay to amplify and classify tau aggregates directly from human brain homogenates. The method utilises two tau substrates, K12 and K11, spanning key aggregation-prone regions, and operates under heparin-free conditions to preserve conformational specificity. Thioflavin T fluorescence maxima, aggregation kinetics (half-times and slopes), and ATR-FTIR spectroscopy were used to distinguish tauopathy subtypes.Our approach enabled the differentiation of eight tauopathy subtypes, including Alzheimer’s disease, Pick disease, progressive supranuclear palsy, corticobasal degeneration, argyrophilic grain disease, FTDP-17 with the N279K mutation, and globular glial tauopathies types II and III. Notably, we achieved subtyping of 4R tauopathies, which have been difficult to resolve to date, through modulation of salt conditions and kinetic profiling. ATR-FTIR analysis confirmed that amplification preserved conformational differences among tau strains. This heparin-free, salt-modulated RT-QuIC platform provides a robust method for the classification of tauopathies directly from brain homogenates. It offers new opportunities for studying tau strain propagation, evaluating therapeutic candidates, and exploring structure-activity relationships in a heparin-free reaction system.

## Introduction

Tau accumulates and forms insoluble amyloid aggregates in a clinically diverse group of neurodegenerative diseases known as tauopathies [1, 2]. Tau is commonly expressed in six isoforms depending on the inclusion of zero, one, or two N-terminal inserts (0N, 1N, 2N) and of three or four microtubule binding repeats (3R, 4R). It has also been reported that in the amyloid state, tau can form over twenty structurally-diverse aggregates that now constitute a structural foundation for the classification of tauopathies [3]. Cryo-electron microscopy studies showed that amyloid structures are shared among patients with similar clinical and neuropathological manifestations [4-6]. These pioneering studies suggest that methods capable of probing the structural properties of tau aggregates in human tissues and, increasingly, in biofluids may prove useful for the molecular diagnosis of neurodegenerative diseases.

The detection and differentiation of protein aggregates has been enabled by the exploitation of their self-propagating properties. The amplification of small amounts of aggregates, referred to as amyloid seeds, is achieved through their ability to grow and multiply in the presence of recombinant monomers of the same protein, including engineered truncations or mutants. The use of amyloid-specific dyes allows for fluorescence-based detection across multiple simultaneous reactions with multi-well plates [7, 8]. One implementation of this technology, called real-time quaking-induced conversion (RT-QuIC), is widely recognized as the protein analogue of the polymerase chain reaction (PCR), wherein singular amyloid proteins (as a singular plasmid target in PCR) can be amplified over a million-fold from human specimens to serve as a diagnostic test for prion diseases [7, 9, 10], synucleinopathies [11-14] and tauopathies [15-18]. While crude brain homogenates are impractical specimens for antemortem diagnosis, they serve as proof-of-concept samples for the development of prototypic seed amplification assays. With improvement in sensitivity and aggregate capture and sampling methods, more clinically relevant specimens like cerebro-spinal fluid (CSF), skin, nasal brushings, urine, and blood can serve as sources of diagnostic aggregates for RT-QuIC assays.

Recently, a range of RT-QuIC assays have been designed to preferentially detect different tau species in distinct tauopathies. These methods achieved selective amplification of 3R tau in Pick disease (PiD) [15], 3R/4R tau in Alzheimer’s disease (AD) and chronic traumatic encephalopathy (CTE) [16], and 4R tau in progressive supranuclear palsy (PSP), corticobasal degeneration (CBD) and some forms of frontotemporal dementia with Parkinsonism linked to chromosome 17 due to *MAPT* mutations (FTDP-17 MAPT) [18]. In these assays, recombinant tau fragments truncated at different points along the microtubule binding region were used to obtain seeding selectivity when combined with brain homogenates or CSF from patients with autopsy-confirmed tauopathies. Interestingly, the 4R tau RT-QuIC assay [18] was developed prior to the discovery of unique structures of 4R tau amyloids.

With the cryo-EM resolution of *ex vivo* 4R tau amyloid structures, we identified a structural basis for the use of trisodium citrate in tau amplification assays, first described in optimisation work and the original 4R tau RT-QuIC [18, 19]. Similarly, we also rationalized the seeding selectivity of the original 4R tau construct, K18, which is truncated 15 residues from the C-terminal-most residues of the insoluble core of *ex vivo* 4R tau fibrils [3, 20]. This substrate choice was made because K18 (4R) and K19 (3R) tau substrates, which are truncated at residue 372 of the canonical tau sequence, were historically used in seed amplification assays due to the inability for AD tau to seed their conversion to mature amyloid [21]. Practically, the knowledge gleaned from the cryo-EM studies on *ex vivo* tau fibrils meant that extending either K18 or K19 to residue 378 could potentially amplify 3R/4R mixed aggregates in addition to their pure 4R or pure 3R amplification capability. Such promiscuity of a substrate was demonstrated when both PiD and AD aggregates could be amplified with an extended 3-repeat tau substrate, K12 tau, that extended to residue 400, beyond the C-terminal most residues of both AD and PiD tau [17]. Divergent thioflavin T (ThT) amplitudes allowed for PiD and AD to be reproducibly discriminated from one another in a combined assay that retained sensitivity and simplified the need for purifying two substrates (K19 and t306-378 could be replaced only with K12).

However, practical methods for subtype-specific classification of tauopathies, particularly among 4R-dominant diseases, remain challenging. As mentioned above, both heparin and a combination of 3R and 4R tau substrates were required to achieve seeding selectivity and sensitivity in our original 4R tau RT-QuIC. To address this problem, here we developed a cryo-EM structure-informed, salt-modulated, heparin-free RT-QuIC platform employing novel tau substrates to enhance conformational specificity and achieve subtype resolution. Building on the success of K12 tau RT-QuIC, and 4R tau RT-QuIC with strain discrimination, we investigated whether an improvement in 4R strain discrimination might be afforded by similarly extending K18 to residue 400. We thus report a combined decision tree framework for selective amplification and structural classification of 3R, 4R, and 3R/4R mixed aggregates with the use of two tau substrates. This decision tree allows for further sub-typing of distinct 4R tauopathies by ThT fluorescence amplitudes and aggregation kinetic analysis in various anionic environments, as previously described [19].

This approach provides a robust and discriminative framework for the molecular classification of tauopathies directly from brain homogenates, offering new opportunities for diagnosis, therapeutic development, and mechanistic investigation of tau strain diversity.

## Results

### K12 and K11 assays produce strain-specific aggregation with sodium salts

Both K12 and K11 tau fragments were expressed in *E. coli* and purified as previously described [17], with the addition of size exclusion chromatography (SEC) to obtain ultra-pure tau (**Figure S1** and **Methods**). K11 tau was designed to include the first, second, third and fourth repeats (R1, R2, R3 and R4), and extends to residue 400 encompassing the entire C-terminal of known *ex vivo* 4R tau amyloids [3], whereas K12 tau comprises repeats R1, R3, and R4, also extending to residue 400 (**Figure S1A**).

We first focused on optimisation of the seeded aggregation of K11 tau RT-QuIC reactions in the presence of sodium salts. Due to the diverse conformation of 4R tau amyloids, we sought reaction conditions that would be permissible to many amyloid conformations and analysed spontaneous aggregation products of K11 in previously-described 1.5 M ionic strength sodium salts [19] (**Figure S2**). The most diverse conformers, indicated by lobations in violin plots, were observed with sodium sulfate (Na_2_SO_4_) and sodium citrate (Na_3_C_6_H_5_O_7_). We then used fold-separation analysis, wherein t_1/2_ control is divided by t_1/2_ positive values, to find the best ionic strength for confident kinetic acceleration of seeded aggregation over spontaneous aggregation. We found that 1.5 M ionic strength reactions achieved the greatest fold separation values throughout reactions, except for those seeded with PSP (**Figure S3**). A final phase of optimisation included substrate concentration, wherein we focused on strain differentiation. By one-way ANOVA analysis, distinctions between further 4R tauopathy sub-types was made possible by conducting the K11 tau RT-QuIC with 4 μM K11 (**Figure S4**). Using these reaction conditions, we observed seeded amplification of K12 tau, indicated by increased ThT fluorescence, in some reactions seeded with brain homogenate dilutions as extreme as 10^−8^ and 10^−7^ for AD and 10^−7^ and 10^−6^ for PiD, respectively, within 80 h (**Figure S5**). In contrast, in reactions seeded with cerebrovascular disease (CVD) lacking tau pathology detectable by immunochistochemistry, we observed spontaneous aggregation of K12 tau at only 10^−4^ – 10^−5^ dilution of brain homogenate, however still delayed compared to reactions seeded wih AD and PiD BH at the same dilutions.

The processing of raw data to ThT maxima and t_1/2_ values used for the classification of tauopathy sub-types in this work are illustrated in **Figure 1**. **Figure 1A,D** shows representative aggregation curves of AD- and PiD BH-seeded K12 tau in the presence of sodium citrate (**Figure 1A;** AD2, PiD6), and GGT type III-, AD-, PSP- and CBD BH-seeded K11 tau reactions in the presence of sodium sulfate (**Figure 1D;** MC24, AD6, 209, 203), each seeded with 1×10^−4^ dilution of brain homogenate. Maxima analysis of **Figure 1A**,**D** yielded the box plots in **Figure 1B**,**E**, respectively, which add statistical backing to strain differences when analysed by one-way ANOVA. **Figure 1C**,**F** represent t_1/2_ values, or time to reaction half-completion in hours, calculated with sigmoidal curve fits, which are stacked vertically beneath their parent curves for visualisation. In good agreement with the previous K12 tau RT-QuIC, we noted reproducible and distinct ThT amplitudes of reactants seeded with AD versus PiD brain homogenates, even in the absence of heparin. This difference was conserved across further brain specimens from additional AD and PiD patients (**Table S1 and Figure S6**).

**Figure 1.**
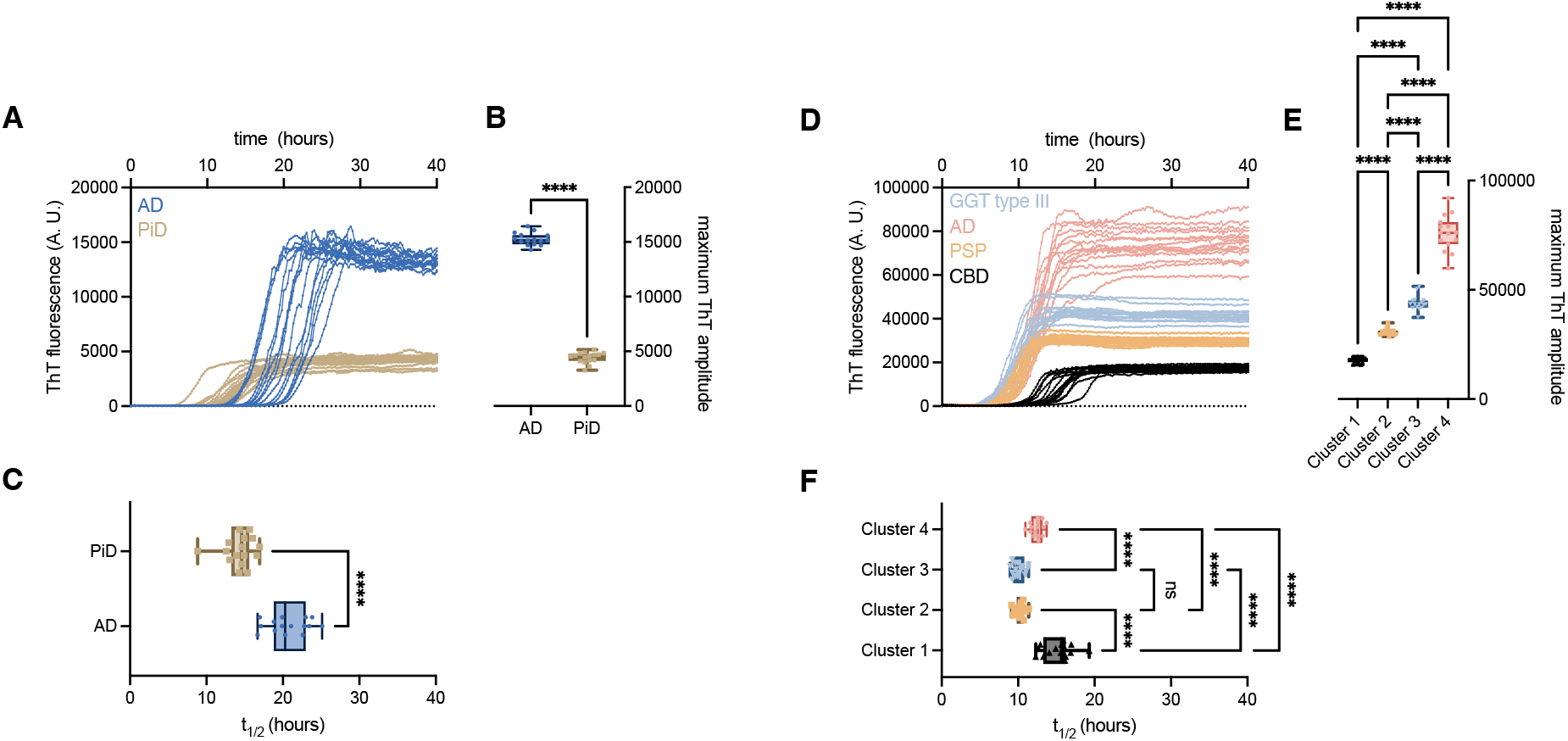
Salt-modulated K12 and K11 RT-QuIC amplification enables differentiation of tauopathy strains from human brain homogenates. **(A-C)** Representative K12 RT-QuIC reactions. **(D-F)** Representative K11 RT-QuIC reactions. **(A**,**D)** Raw ThT fluorescence traces from sixteen replicate reactions seeded with individual brain homogenates. (**B**,**E**) ThT fluorescence maxima corresponding to curves in (**A**) and (**D**), illustrating clustering by disease subtype. **(C**,**F)** Aggregation half-times (t_1/2_) calculated from sigmoidal curve fits of the raw traces. Statistical significance was assessed by one-way ANOVA; ^****^ p < 0.0001; ^***^ p < 0.001; ns, not significant. In the box plots, boxes represent the interquartile range, horizontal lines indicate the median, and whiskers denote the full data range. Brain samples used in this figure include AD2, PiD6, 203, 209 and MC21.

4R BH-seeded K11 reactions produced four basic clusters when ThT maxima were analysed (**Figure 1E**). The example data in **Figure 1** illustrate reactions seeded by CBD (cluster 1), PSP (cluster 2), GGT type III (cluster 3), and AD (cluster 4). Divergent CBD and PSP amplitudes were in good agreement with those observed in the original 4R tau RT-QuIC assay [18] as well as a case report of an un-classified tauopathy which used K11 tau [22]. New to this work is the addition of AD-seeded 4R aggregation which occupied its own ThT maximum cluster. Additionally, in cluster III, GGT type III and type II seeded K11 into a conformer that exhibited a distinct ThT fluorescence maximum. Simple kinetic analysis of half-times (t_1/2_) of reactions supported previous findings where CBD-seeded K11 reactions were markedly different from those seeded with PSP (**Figure 1F**). These differences were conserved across further brain specimens from additional patients (**Table S1 and Figure S7**).

### Anionic salts differentiate 4R tau amyloids in the absence of heparin

We then sought to investigate if further sub-classification of 4R tauopathy amyloids could be achieved with K11 tau in the absence of heparin. We previously observed up to million-fold enhancement of sensitivity of 4R tau detection in RT-QuIC assays with the use of strongly-hydrated anions [19]. From cryo-EM reconstructions of *ex vivo* 4R tau fibrils, it appears that an anionic density is present in the core of some filaments, serving to neutralise lysine and histidine residues [3, 23]. Because of this, we sought to investigate whether 4R aggregates could be sub-classified by assaying in distinct sodium salts. **Figure 2** shows ThT maxima of K11 reactions seeded with individual brain homogenates in (**A**) sodium sulfate and (**B**) sodium citrate. Each violin represents a different patient brain homogenate. Each point represents one of 16 replicate reactions. We observed again the four basic clusters observed in **Figure 1**. CBD-, argyrophilic grain disease (AGD)- and N279K-seeded K11 reactions resulted in low ThT amplitudes highlighted in grey background. PSP-seeded reactions occupied the same cluster 2 range whether seeded in the presence of sulfate or citrate. GGT type II and III BH-seeded K11 reactions occupied a unique cluster 3 when in the presence of sulfate, however GGT type III reactions shifted to match cluster 2-like ThT maxima in the presence of sodium citrate, representing a method for GGT sub-classification by salt modulation. 3R/4R AD BH-seeded K11 reactions represent a singular cluster in both salts, distinct from all pure 4R BH-seeded reactions. This difference was conserved across further brain specimens from additional patients (**Table S1 and Figures S7**).

**Figure 2.**
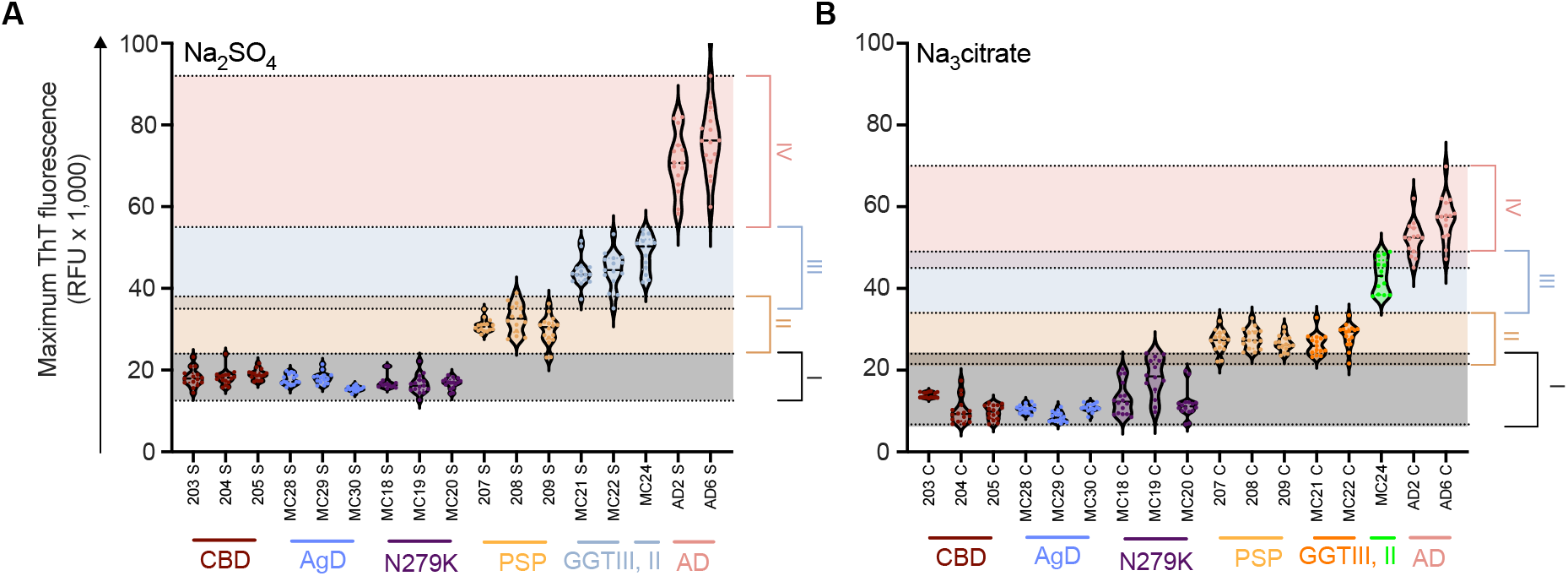
Salt-modulated K11 RT-QuIC amplification differentiates 4R tauopathy subtypes by ThT fluorescence maxima. (**A**,**B**) ThT fluorescence maxima from sixteen replicate K11 RT-QuIC reactions seeded with individual brain homogenates are shown for reactions performed in 500 mM sodium sulfate (**A**) and 250 mM sodium citrate (**B**). Distinct clustering of tauopathy subtypes is observed under both conditions, corresponding to CBD (cluster 1), PSP (cluster 2), GGT type II and III (cluster 3), and AD (cluster 4). Notably, GGT type III-seeded reactions shift from cluster 3 toward cluster 2 under sodium citrate conditions, enabling differentiation of GGT type II and III. AD-seeded reactions consistently form a distinct high-fluorescence cluster across both salt conditions. Shaded areas represent approximate ThT fluorescence ranges defining each cluster. Each point represents a single replicate reaction.

### Kinetic differentiation of CBD, AGD, and N279K tauopathies

Previously, our 4R tau RT-QuIC assay was unable to differentiate CBD from N279K FTDP and AGD [18]. To address this problem, we conducted further kinetic analysis of the reactions comprising cluster I. **Figure 3A** shows average +SD of 16 replicate reactions staggered ∼15,000 fluorescence units for comparison. **Figure 3B** presents the t_1/2_analysis performed as described above and **Figure 3C** shows the Hill slope analysis of curve-fitted simple sigmoids. We observed that, beyond ThT maxima clustering these reactions together, there was a statistically significant difference in half-times between the three unique disease-seeded K11 reactions. Such kinetic analysis with K11 was used with ThT maxima analysis previously to sub-classify an unknown case into a yet unclassified tauopathy which showed neuropathologic, clinical, and biochemical features that were intermediate between PSP and CBD [22]. In addition to divergent t_1/2_values, we noted significantly steeper Hill slope analysis in N279K FTDP-seeded K11 reactions compared to CBD and AGD, which may reflect faster filament elongation or fragmentation rates, or a combination of the two. These findings were further validated by performing a seed dilution experiment to assess whether the observed differences were a result of seed dose effects (**Figure S8**). A significant delay of t_1/2_aggregation values of CBD-seeded K11 reactions in sodium citrate, relative to sodium sulfate is observed while there is no such effect on AGD-seeded reactions.

**Figure 3.**
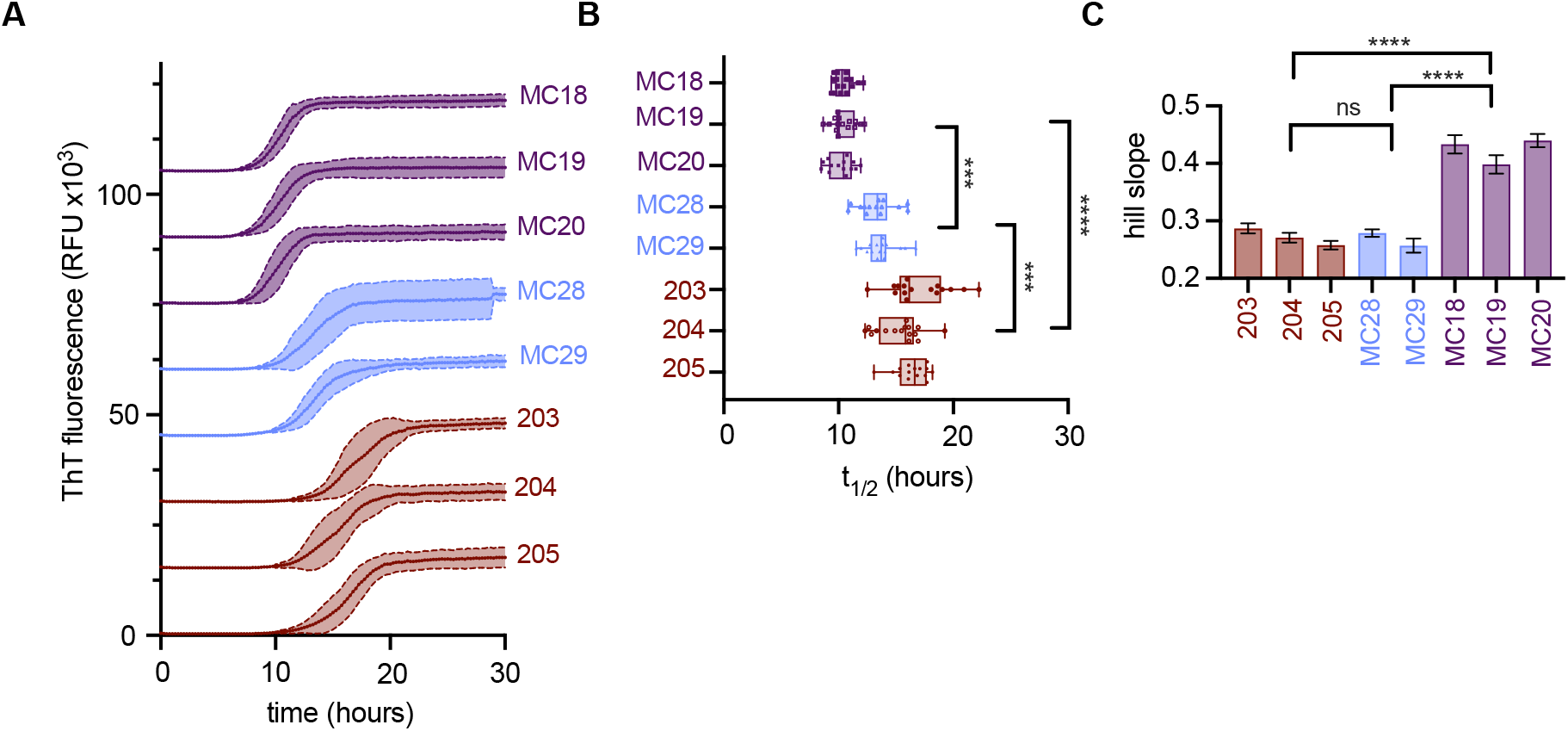
Kinetic analysis of K11 RT-QuIC reactions enables sub-classification of tauopathies within cluster 1. **(A)** Representative ThT fluorescence traces from sixteen replicate K11 RT-QuIC reactions seeded with brain homogenates from patients with FTDP-17 N279K, AGD, and CBD. Curves are staggered by 15,000 relative fluorescence units for visualization. **(B)** Aggregation half-times (t_1/2_) derived from sigmoidal fits of the fluorescence curves reveal significant differences between the three disease groups. **(C)** Hill slope analysis of the fitted curves shows that N279K-seeded reactions exhibit steeper slopes compared to AGD- and CBD-seeded reactions, indicating faster aggregation kinetics. Statistical comparisons were performed using one-way ANOVA; ^****^ p < 0.0001, ^***^ p < 0.001. In box plots, boxes represent interquartile ranges, horizontal lines indicate medians, and whiskers denote full data ranges.

### ATR-FTIR analysis of BH-seeded K12 and K11 reactions indicates the amplification of unique structures

We observed previously with tau RT-QuIC assays that the amplified products, when recovered an analysed by ATR-FTIR, show unique β-sheet vibrational modes that correlate with ThT amplitude differences, suggesting amplification of divergent amyloid structures [17, 18]. **Figure 4** reports second derivative ATR-FTIR spectra of (top) K11 reaction products and (bottom) K12 reaction products in the presence of heparin (dashed lines) and absence of heparin (solid lines). **Figure 4A**,**D** represent the FTIR spectrum region comprised largely of β overtones and turn vibrations, **Figure 4B**,**E** represent main β-sheet vibrational modes, and **Figure 4C**,**F** show the tyrosine vibrational mode. In good agreement with the original 4R tau RT-QuIC which used K18 and K19 tau substrates, we observed divergent FTIR spectra relative to the original ThT maxima-defined clusters. Minimal vibrational differences are observed in the presence or absence of 40 μM heparin. Vertical dotted lines represent previously-reported vibrational modes for CBD (1629 and 1667 cm^-1^), PSP (1676 and 1667 cm ^-1^), AD (3R, 1630 and 1618 cm ^-1^), PiD (1625 and 1633 cm^-1^). Of note, main β-sheet vibrational modes of AD-seeded K11 and K12 products differ, suggesting distinct propagated structures. We also report a novel propagated 4R tau RT-QuIC structure, GGT type II- and GGT type III-seeded (cluster 3, sodium sulfate) K11 reaction products which show unique β-vibrational overtones 1660 and 1672 cm^-1^.

**Figure 4.**
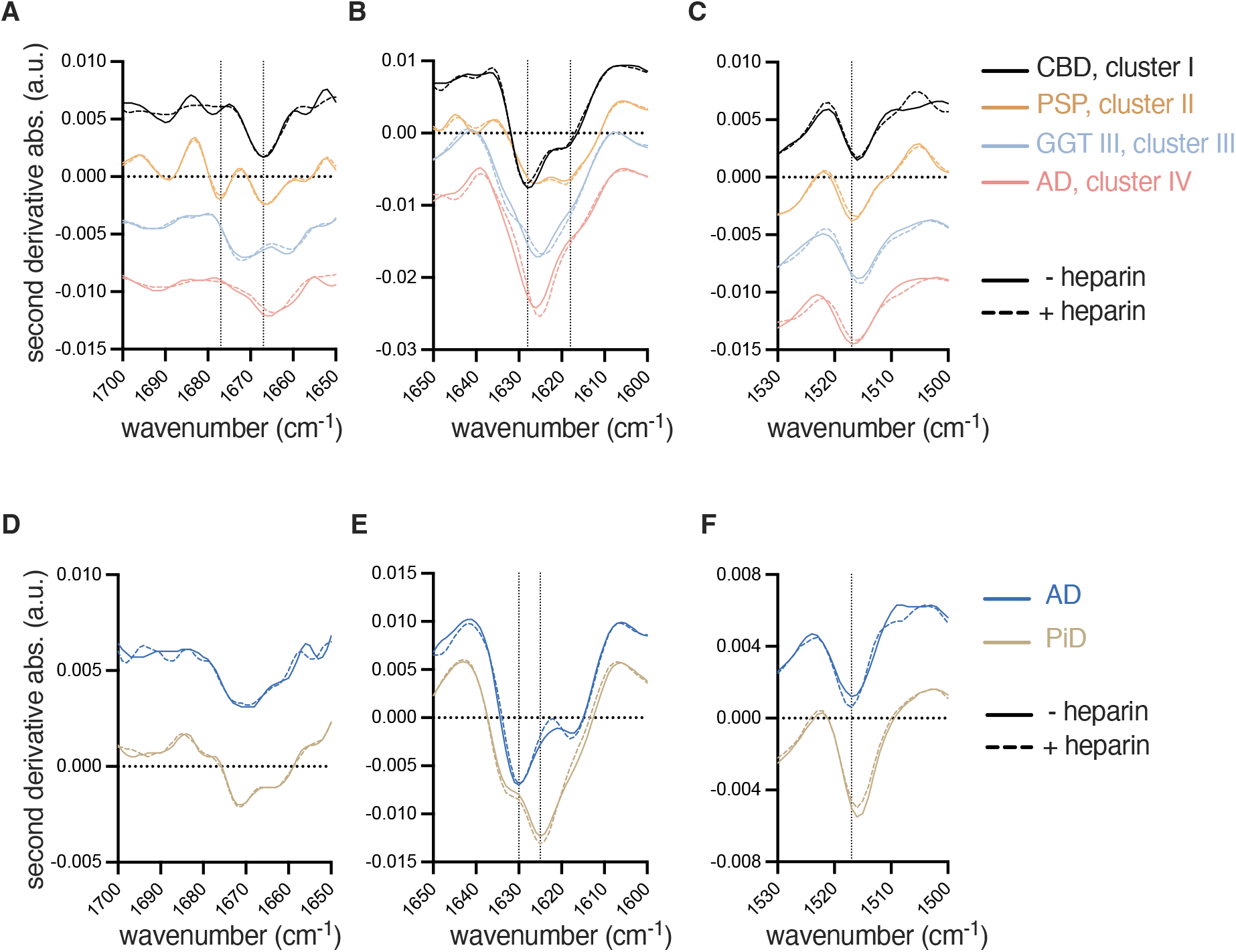
ATR-FTIR spectroscopy confirms strain-specific structural amplification by salt-modulated K11 and K12 RT-QuIC. **(A-F)** Second-derivative ATR-FTIR spectra of pooled RT-QuIC reaction products are shown for K11 (A-C) and K12 reactions (D-F). Solid lines represent heparin-free conditions using sodium salts; dashed lines represent reactions performed in the presence of heparin. (**A**,**D)** Overtones and turn vibration regions. **(B**,**E)** Main β-sheet vibrational modes (1600– 1700 cm^−1^), highlighting strain-specific spectral differences. **(C**,**F)** Tyrosine vibrational modes. Distinct β-sheet vibrational profiles correspond to different tauopathy seeds, consistent with ThT fluorescence clustering. Minimal spectral differences are observed between reactions performed with and without heparin, supporting structural fidelity under salt-modulated conditions. Notably, GGT-seeded K11 products exhibit a unique β-sheet signature distinct from other 4R tauopathies.

### Stepwise classification of tauopathies using K12 and K11 RT-QuIC assays

Based on the results described above, we systematically classify tauopathies with a stepwise approach integrating K12 and K11 RT-QuIC assays with cofactor modulation (**Figure 5**). First, we applied the K12 RT-QuIC assay to assess seeding activity. Samples exhibiting seeding relative to the control were classified as either AD or PiD, which were further distinguished based on ThT fluorescence amplitudes. In contrast, samples that did not show seeding in the K12 assay were subjected to the K11 RT-QuIC assay in the presence of sodium sulfate to determine whether they belonged to a 4R tauopathy. Fluorescence amplitude analysis in the K11 RT-QuIC assay revealed four distinct clusters. With ThT fluorescence maxima analysis alone, a sample with cluster 1 ThT maximum cannot be distinguished between CBD, AGD, or N279K FTDP. Further sub-classification of these unknowns takes the form of t_1/2_analysis and Hill slope analysis where such brains can be terminally differentiated. An unknown sample that shows seeding with ThT amplitude in the range of 25 – 40 k RFU must be PSP. Those with amplitude between 40 – 55 k could be either GGT type II or III, however with assaying in sodium citrate, following along the dichotomous key, a relaxation of ThT fluorescence into cluster II occurs with GGT type III only, which serves to terminally differentiate these diseases. Cluster IV is comprised only of 3R/4R mixed diseases AD. While the 4R tau RT-QuIC assay is not required to diagnose these, we observed no overlap of additional 4R-only specimens in this ThT fluorescence range, which can add confidence when performing K12 and K11 assays in concert.

**Figure 5.**
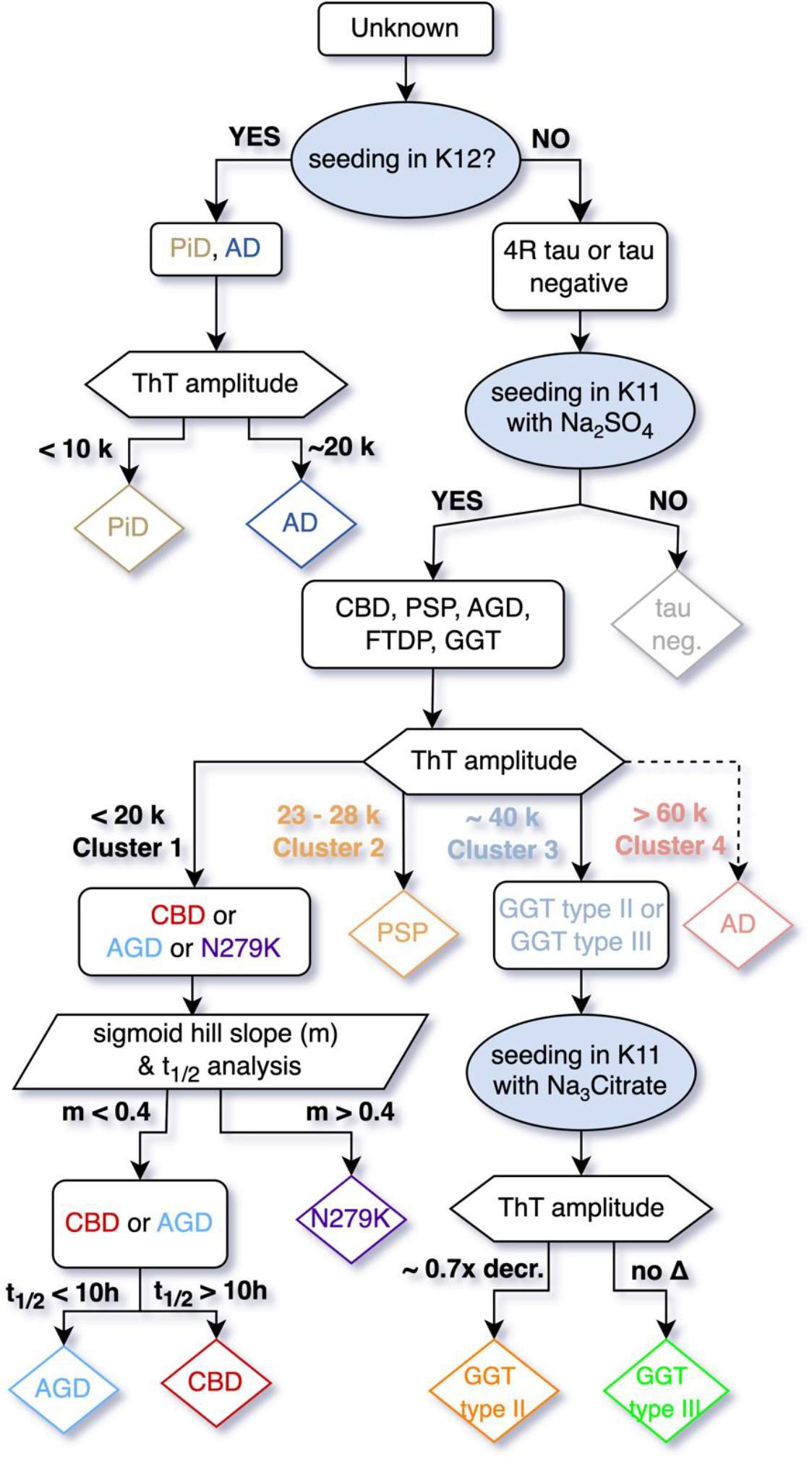
Stepwise classification of tauopathies using salt-modulated K12 and K11 RT-QuIC assays. A decision tree integrating K12 and K11 RT-QuIC results enables systematic classification of tauopathies from human brain homogenates. Samples are first assessed with K12 RT-QuIC, with positive seeding distinguishing Alzheimer’s disease (AD) and Pick disease (PiD) based on ThT fluorescence maxima. K12-negative samples are subjected to K11 RT-QuIC in 500 mM sodium sulfate. ThT maxima analysis clusters samples into four groups: Cluster 1 (CBD, AGD, N279K FTDP-17), Cluster 2 (PSP), Cluster 3 (GGT type II and III), and Cluster 4 (3R/4R tauopathies, predominantly AD). Further discrimination within clusters is achieved by kinetic analysis (half-time and Hill slope) and by assessing shifts in ThT maxima under 250 mM sodium citrate conditions. This combined approach enables high-resolution, strain-level differentiation of major tauopathy subtypes.

## Discussion

With the advent of cryo-EM analysis of *ex vivo* amyloids of neurodegenerative disease, we are edging towards structure-based disease classification [3]. The discovery of identical amyloid structures in patients with both preclinical and neuropathologic diagnosis of tauopathy underscores the potential of amyloid strain detection as a diagnostic tool [3, 4]. To capitalize on these insights, tau RT-QuIC assays have been reported for the detection of aggregates based on the use of different tau substrates, 3R [15], 3R/4R mixed [16], 4R [18], and a combination assay that could amplify 3R and 3R/4R mixed aggregates while preserving strain discrimination [17]. Inspection of the cryo-EM models reveals an extensive anionic network within the 4R core, explaining the empirical efficacy of polyvalent anions like trisodium citrate [17-19], as well as inform the rational design of tau substrates for assay optimisation and simplification [17]. Guided by these structural principles, we now established a framework that use only two tau substrates to amplify eight tau subtypes (**Figure 5**).

We introduced two main advances in this work to enhance the discrimination of tau aggregate conformers in the tau RT-QuIC assay. First, we extended the tau fragment to residue 400 to improve the subtyping within the 4R tauopathy spectrum, surpassing the previous K18-based assay [18]. Additionally, since heparin-induced tau amyloids have failed to recapitulate disease-associated structures [24], we removed the heparin co-factor by incorporating 250 mM sodium citrate and 500 mM sodium sulfate, thus enabled heparin-free amplification of tau aggregates from brain homogenates. Additionally, purification of K12 and K11 using size exclusion chromatography, beyond a standard His-trap, yielded highly pure monomeric tau, improving the clustering of ThT maxima and refining 4R strain subtyping (**Figure S1**).

The 4R tau RT-QuIC assay previously achieved selectivity for solely 4R tauopathies by truncating the K18 substrate at residue 372, just before the ^373^THKLTF^378^ motif critical for 3R/4R mixed aggregation [18]. Extending the tau fragment to residue 400 (K11) introduced seeding activity for AD but retained the ability to distinguish tauopathies via ThT fluorescence maxima, similar to how K12 enabled PiD-AD discrimination [17]. The K11 assay replicated the previously reported ThT maxima differences between PSP and CBD [22], reinforcing their classification as distinct 4R tauopathies. Prior to its formal definition, some cases of GGT were diagnosed as atypical PSP [25]. However, with GGT being recognised as a separate neuropathological entity, the K11 RT-QuIC assay achieved discrimination of PSP from GGT, with sodium sulfate aiding in GGT differentiation. Further subtyping of GGT type II and GGT type III was achieved through selective modulation with sodium sulfate and sodium citrate, with GGT type III shifting to cluster 2-like ThT maxima in the presence of sodium citrate, enabling GGT sub-classification.

Fluorescence intensity clustering of AGD, CBD, and N279K FTDP showed similar ThT maxima across sodium sulfate and sodium citrate, but kinetic analysis refined their classification. Differences in t_1/2_ and Hill slope values highlighted distinct aggregation rates, confirmed by seed dilution experiments, ruling out seed concentration effects. Nevertheless, the similarities observed in the Hill slope and ThT maxima between AGD and CBD are consistent with their classification into the same 4R tau subtype, as both diseases share the characteristic structural feature of the C-terminal domain packing against part of R2 [3].

In addition to the observed differences in ThT fluorescence maxima, ATR-FTIR spectroscopy has proven effective in distinguishing PiD-seeded from 3R/4R-seeded RT-QuIC reactions [17], as well as discriminating CBD from PSP [18]. These findings point to the presence of distinct conformational signatures in the filaments amplified by the assay. Consistent with the ThT fluorescence profiles, ATR-FTIR revealed four distinct spectral clusters among the K11 reaction products, as well as clear differences between AD- and PiD-seeded K12 reactions. The most prominent spectral differences were localised within the 1618-1633 cm^−1^ region, a range typically associated with β-sheet backbone vibrations [26, 27]. The distinct β-sheet-associated vibrational modes detected in AD-seeded K11 versus K12 reactions suggest the templated propagation of structurally divergent assemblies. Nevertheless, the focus of this study is to detect and differentiate the seeding-dependent reaction products, rather than to resolve the precise structural details of the filaments they produce.

We note that while our assays robustly differentiate tau strains based on seeding activity and aggregate conformation, further studies combining RT-QuIC with high-resolution structural methods such as cryo-EM or nanoscale FTIR could refine our understanding of the precise conformational fingerprints associated with each tauopathy.

In conclusion, compared to prior tau RT-QuIC assays, which were limited by substrate promiscuity and reliance on non-physiological cofactors like heparin, our method enables high-resolution tauopathy discrimination under conditions that better preserve native conformational properties. The heparin-free K11 and K12 RT-QuIC assays are being applied for use in a variety of settings including strain differentiation of complex neuropathologic cases [22], for the study of seeding activity in mouse models of tauopathy [28], and for conformation-specific small molecule inhibitor discovery [29]. The capacity to differentiate 4R tauopathies at the subtype level directly from brain tissue could serve as a foundation for developing biomarker assays for early, minimally invasive diagnosis from peripheral samples such as CSF or nasal brushings.

## Materials and Methods

### Neuropathology and compliance with ethical standards

*Post-mortem* frozen frontal cortex brain samples from cases confirmed neuropathologically were supplied by the Dementia Laboratory in the Department of Pathology at Indiana University School of Medicine or the Mayo Clinic brain bank for neurodegenerative disorders. Fifteen samples were provided by Drs Shunsuke Koga and Dennis Dickson, Mayo Clinic, Jacksonville Florida; they included progressive supranuclear palsy (PSP x 3), corticobasal degeneration (CBD x 3), argyrophilic grain disease (AGD x 3), glial globular tauopathy (GGT x 3) type II and GGT type III, frontotemporal dementia with parkinsonism associated with chromosome 17 with N279K mutation (FTDP-17 N279K x 3). The remaining seven samples were provided by Dr Bernardino Ghetti, Indiana University school of medicine and included cerebrovascular disease (CVD x 1), Pick disease (PiD x 3), and Alzheimer’s disease (AD x 3). These brain tissue specimens were obtained *post-mortem* with written informed consent from patients or their legal representatives given to the Indiana University School of Medicine or the Mayo Clinic. No additional ethical permission was needed because the samples were taken from deceased, deidentified, consenting individuals. Such autopsy samples are considered exempt from human subject research by the State of Indiana and the Mayo Clinic Institutional Review Board. **Table S1** provides a detailed summary of the demographics, clinical diagnoses, the institution of origin for each case, and the biochemical and structural classification resulted from this study. In summary, one half of each brain was preserved in formalin while the other half was frozen. Diagnoses of the preserved tissues were carried out using established immunohistochemical staining techniques [16, 30]. For RT-QuIC analysis, brain tissue was homogenized to yield a 10% (w/v) solution in ice-cold phosphate-buffered saline (PBS) + protease inhibitor cocktail buffer at pH 7.4 using a multi-bead shocker (Fisher). Supernatants were collected from a 2-min centrifugation step at 2000×g and stored at −80 °C until use.

### Purification of recombinant K12 and K11 tau

A cloning cassette encoding the K12 tau fragment (residues 244–275 and 306–400 of full-length human tau) with an alanine mutation at residue 322, and the K11 tau fragment (residues 244-400 and 306-400) with alanine mutations at residues 291 and 322, was inserted between the *NdeI* and *XhoI* sites of the pET28 bacterial vector, resulting in an N-terminal histidine tag. Cysteine mutations were introduced to prevent disulfide-mediated dimerisation. K11 and K12 tau constructs were expressed in *E. coli* BL21(DE3) cells using an overnight autoinduction protocol with slight modifications [15, 17]. Cells were harvested by centrifugation (4,000×*g*, 20 min, 4 °C) and resuspended in buffer A (40 mM Tris, 400 mM NaCl, 5 mM imidazole, pH 7.4) supplemented with a cOmplete^™^ EDTA-free protease inhibitor cocktail (10 mL/g pellet). Lysates were centrifuged (14,000×*g*, 35 min, 4 °C) and filtered through 0.45 µm pore filters before loading onto a HiTrap 5 mL column (GE Healthcare, 17–5255-01). After washing with buffer A, elution was performed over a step gradient of 10%, 40%, and 100% of buffer B (40 mM Tris, 400 mM NaCl, 300 mM imidazole, pH 7.4) over 30 column volumes. Fractions were analysed by SDS-PAGE, pooled, and precipitated in 75% acetone overnight at 4 °C. Precipitates were centrifuged (4,700 × *g*, 20 min), acetone was decanted, and pellets were dissolved in 4 M guanidine-HCl (GdnHCl). The solution was purified using a Superdex 75 column (26 × 600 mm) equilibrated in PBS. Tau fractions were analsed by SDS-PAGE, and pure monomer fractions were pooled, aliquoted, lyophilised, and stored at –80 °C.

### K12 and K11 RT-QuIC assays

The K12 and K11 RT-QuIC assays follow a modified version of the protocols previously published [17, 18]. NaF and heparin were replaced with 250 mM sodium citrate and 500 mM sodium sulfate. Lyophilised K12 and K11 tau aliquots were resuspended in water, filtered with a 100 kDa spin column filter (Pall, OD100C35) to remove any possible aggregates. Brain homogenates were thawed from storage at 10% w/v in ice cold PBS and diluted in 10x N-2 Supplement (Gibco) to the designated concentration; assays were performed at 1×10^−4^, 1×10^−5^, and 1×10^−6^ concentration of brain homogenate. Previously, dilution was performed using 0.53% tau-free mouse brain homogenate (hTau KO). Here, N2 supplement was increased 10-fold compared to previously published tau RT-QuIC assays as a dilution and reaction matrix; we found the use of 10X N-2 critical for preventing spontaneous aggregation of the tau fragments (reaction matrix) as well as providing a preventing loss of aggregates on plastic surfaces (dilution matrix). 1 μL of each brain homogenate was first diluted to 10-fold with 10x N2 supplement (Gibco) in DI water. 0.5 μL of diluted brain homogenate solution was again 100-fold diluted to seed 50 μL of reactions in the presence of 2 or 4 μM K12/K11, 10 μM ThT, 250 mM Na_3_C_6_H_5_O_7_ or 500 mM Na_2_SO_4_, and 10 mM HEPES buffered at pH 7.4. In contrast to previous tau RT-QuIC methodology wherein reaction mix was added to microwell plates and then seeded directly with microliter amounts of brain homogenate, reactions were seeded directly into the reaction mixes and mixed thoroughly, and then split into wells of 384-well optical bottom plates (Nunc, 236366) and sealed with tape. The plate was incubated at 37 °C in a plate reader (BMG Labtech FLUOstar Omega) with periodic shaking: 60 seconds of orbital shaking at 500 rpm followed by 60 seconds of no shaking. Periodic ThT fluorescence readings every 15 min from the bottom were recorded using 450-10 nm excitation and 480-10 nm emission. Instrument gains were kept between 800-1000 relative fluorescence units (rfu) to avoid detector saturation.

### RT-QuIC data processing and statistical analysis

Data processing was performed with GraphPad Prism version 10.1. Half-time (t_1/2_) analysis was performed by fitting dose-dependent sigmoid curves to individual raw ThT fluorescence plots wherein log_EC50_output values are reported as half-times. ThT maxima were obtained by ‘area under the curve’ function in Prism wherein maximum values are reported. Hill slopes were reported from maximum slopes of fitted dose-dependent sigmoid curves, averaged over 16 replicate reactions where error bars represent one standard deviation of fitted Hill slopes. Analysis of statistically significant differences between ThT maxima and t_1/2_ values was conducted by one-way ANOVA with multiple comparisons of means. For all comparisons herein: ^****^ p < 0.0001, ^***^ p < 0.001, ^**^ p < 0.01, ns nonsignificant.

### FTIR analysis of K12 and K11 RT-QuIC products

3–16 replicate K12 or K11 tau RT-QuIC reactions seeded at at a 10^−4^ dilution of brain homogenate were pooled by scraping from the bottom of the 384-well plates with a pipette tip and transferred for analysis. These reactions were stopped when ThT fluorescence reached its plateau. The combined samples were then centrifuged at 21,000×g for 10 min at 4 °C, the supernatant was removed and 5 µL of reaction buffer was used to resuspend the concentrated pellet. A 1.5 µL aliquot of the sample was applied to a Perkin Elmer Spectrum 100 FTIR with a diamond crystal ATR attachment. Samples were air-dried until the 3400 cm^−1^ H2O band stopped fluctuating. 100 scans were taken and averaged from 800 to 4000 cm^−1^ with a 4 cm^−1^ resolution for each sample, using strong apodization, and both the sample and electronic chambers were continuously purged with dry air or nitrogen. The spectra were normalized to the amide I intensity (1600-1700 cm^−1^ range) and second derivatives were calculated using nine points for slope analysis.

## Data Availability

All data produced in the present work are contained in the manuscript

## Acknowledgements

We acknowledge the support of the Centre for Misfolding Diseases (A.S. and M.V.); MM would like to acknowledge the NIH-Cambridge scholars programme and the Cambridge Trust. This work was supported by the Department of Pathology and Laboratory Medicine, Indiana University School of Medicine (R01AG080001 and P30AG072976 to B.G.).

