## Supplementary Information for "Classification of Tauopathies from Human Brain Homogenates through Salt-Modulated Tau Amplification"

**Supplementary Table 1**. Clinical, neuropathologic, and RT-QuIC diagnostic data of samples used in this study

| | clinical and neuropathological data | | | | | | | | molecular diagnostic data | | | | | | | | | | | | | |
|---|---|---|---|---|---|---|---|---|---|---|---|---|---|---|---|---|---|---|---|---|---|---|
| Assay | Tauopathy Class | Clinical Dx | 1° (2°, 3°, 4°) Neuropath. Dx | Brain Region | Sex | AOD | PMI | brain weight (g) | RT-QuIC | | | Heparin-free FTIR bands (Present / Absent) | | | | | | | | | | |
| | | | | | | | | | ThT amp (A.U.) | t1/2 avg (h) | hill slope | 1676 | 1672 | 1667 | 1661 | 1631 | 1629 | 1628 | 1625 | 1618 | 1617 | tyrosine |
| K12 (3R) | 3R/4R | AD | sAD 2† | F | F | 80-84 | 26 | 1007 | 15264 | 20.69 | | | | | | | | | | | | 1517 |
| | | AD | sAD 5† | F | M | 70-74 | 2 | | 16595 | 13.87 | | | | | | | | | | | | 1517 |
| | | AD | sAD 6† | F | F | 80-84 | 33 | 851 | 16884 | 17.42 | | | | | | | | | | | | 1517 |
| | 3R | PiD | PiD 5 (CVD)† | F | M | 65-70 | 13 | | 6566 | 15.26 | | | | | | | | | | | | 1515 |
| | | PiD | PiD 6 (CVD)† | F | F | 75-79 | 9 | 891 | 4416 | 14.35 | | | | | | | | | | | | 1515 |
| | | PiD | PiD 7 (SC, CVD)† | F | F | 70-74 | 24 | 993 | 6837 | 12.46 | | | | | | | | | | | | 1515 |
| K11 (4R) | 3R/4R | AD | sAD 2† | F | F | 80-84 | 26 | 1007 | 70774 | 14.72 | | | | | | | | | | | | 1517 |
| | | AD | sAD 6† | F | F | 80-84 | 33 | 851 | 77590 | 12.31 | | | | | | | | | | | | 1517 |
| | 4R | CBD | 203 (CBD)* | F | M | 60-64 | | | 18190 | 16.42 | 0.287 | | | | | | | | | | | 1516 |
| | | CBD | 204 (CBD)* | F | M | 70-74 | | | 18188 | 15.35 | 0.271 | | | | | | | | | | | 1516 |
| | | CBD | 205 (CBD)* | F | M | 70-74 | | | 18890 | 17.15 | 0.257 | | | | | | | | | | | 1516 |
| | | PSP | 207 (PSP)* | F | F | 75-79 | | | 39833 | 10.18 | | | | | | | | | | | | 1517 |
| | | PSP | 208 (PSP)* | F | F | 65-69 | | | 32665 | 15.11 | | | | | | | | | | | | 1517 |
| | | PSP | 209 (PSP)* | F | M | 65-69 | | | 29698 | 13.17 | | | | | | | | | | | | 1517 |
| | | PPND | MC18 (N279K FTD)* | SFG | F | 50-54 | 18 | 1040 | 16990 | 10.21 | 0.433 | | | | | | | | | | | 1516 |
| | | PPND | MC19 (N279K FTD)* | SFG | M | 40-44 | n/a | 1000 | 16522 | 10.43 | 0.398 | | | | | | | | | | | 1516 |
| | | PPND | MC20 (N279K FTD)* | SFG | F | 45-49 | 19 | 1140 | 16748 | 10.41 | 0.44 | | | | | | | | | | | 1516 |
| | | PSP + dementia | MC21 (GGT type III)* | F | F | 65-69 | 18 | 1001 | 43916 | 9.93 | | | | | | | | | | | | 1517 |
| | | PSP | MC22 (GGT type III)* | F | F | 65-69 | 20 | 1010 | 44392 | 9.83 | | | | | | | | | | | | 1517 |
| | | AD, PSP | MC24 (GGT type II)* | F | F | 80-84 | 17 | 990 | 48687 | 11.77 | | | | | | | | | | | | 1516 |
| | | | MC28 (AGD)* | F | F | 80-84 | 19 | 1049 | 17698 | 13.74 | 0.278 | | | | | | | | | | | 1516 |
| | | | MC29 (AGD)* | F | M | 90-94 | 2 | 1175 | 18086 | 13.33 | 0.257 | | | | | | | | | | | 1516 |
| | | | MC30 (AGD)* | F | M | 85-89 | 11 | 1330 | 16408 | 13.44 | 0.31 | | | | | | | | | | | 1516 |
| | IHC no tau | | CVD 1† | F | F | 50-54 | 19 | 1192 | | 25.3 (4R), 41.2 (3R) | | | | | | | | | | | | |

Neuropathological diagnoses were provided for the indicated cases (†) by Dr. Bernardino Ghetti (Indiana University), (*) Drs. Shunsuke Koga and Dennis Dickson (Mayo Clinic)

AGD, Argyrophilic grain disease; AD, Alzheimer Disease; CBD, Corticobasal Degeneration; CVD, Cerebrovascular Disease; GGT, Globular glial tauopathy; N279K FTD, Frontotemporal dementia with N279K MAPT mutation; PiD, Pick's disease; PSP, progressive supranuclear palsy; PPND, pallidopontonigral degeneration; AOD, age at death; PMI, postmortem interval; F, frontal lobe; SFG, superior frontal gyrus

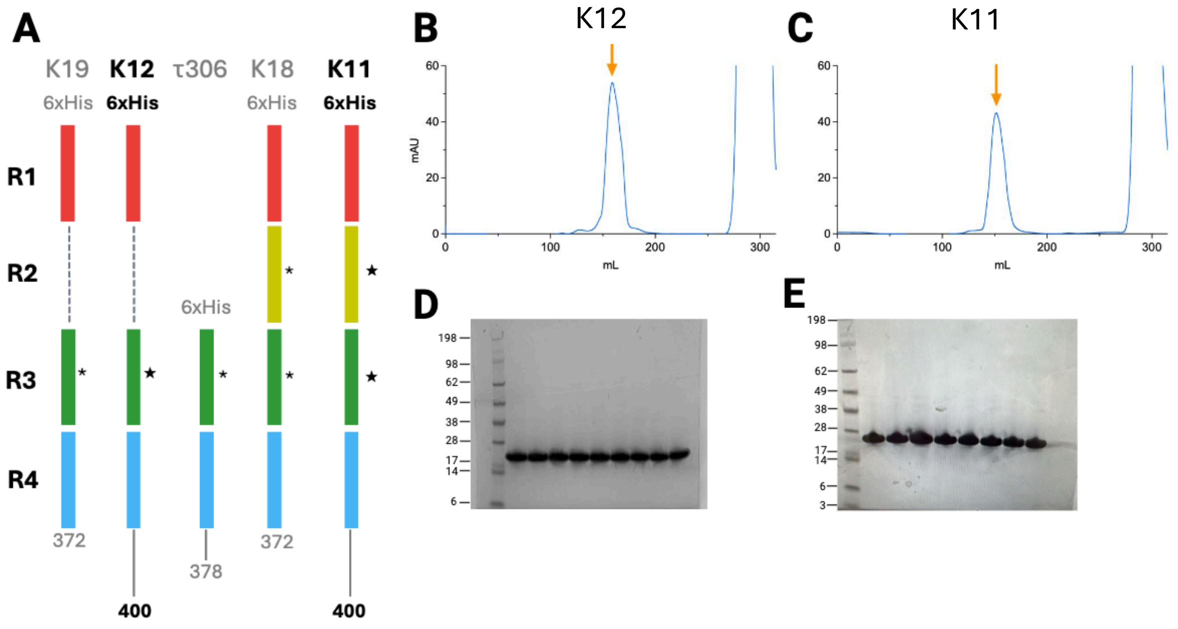


**Figure S1. Sequences and purification of K11 and K12 tau.** (**A**) Schematics of truncated and mutated K11 and K12 RT-QuIC substrates relative to the previously

used RT-QuIC substrates. (**B**) Size exclusion chromatography (SEC) of K12. (**C**) SEC of K11. (**D**) Coomassie-stained SDS-PAGE analysis of K12 tau after SEC. (**E**) SDS-PAGE of K11 tau after SEC.

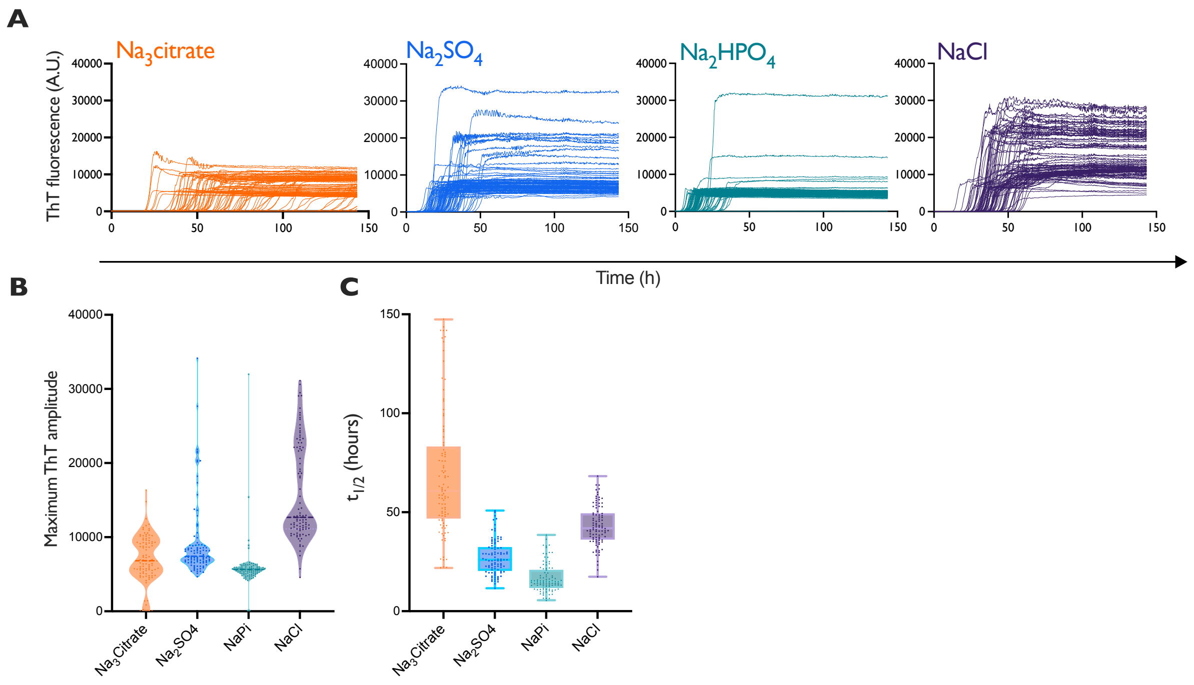


**Figure S2. Optimisation of K11 tau RT-QuIC: selection of sodium salts. (A)** ThT fluorescence traces from 192 replicate spontaneous K11 substrate reactions performed at an ionic strength of 1.5 M in various sodium salts. **(B)** Violin plots of ThT fluorescence maxima illustrating conformational clustering across different salts. **(C)** Box-and-whisker plots showing the distribution of aggregation half-times ($t_{1/2}$) corresponding to the reactions in (**A**). boxes represent inter-quartile range, whiskers represent outer quartiles.

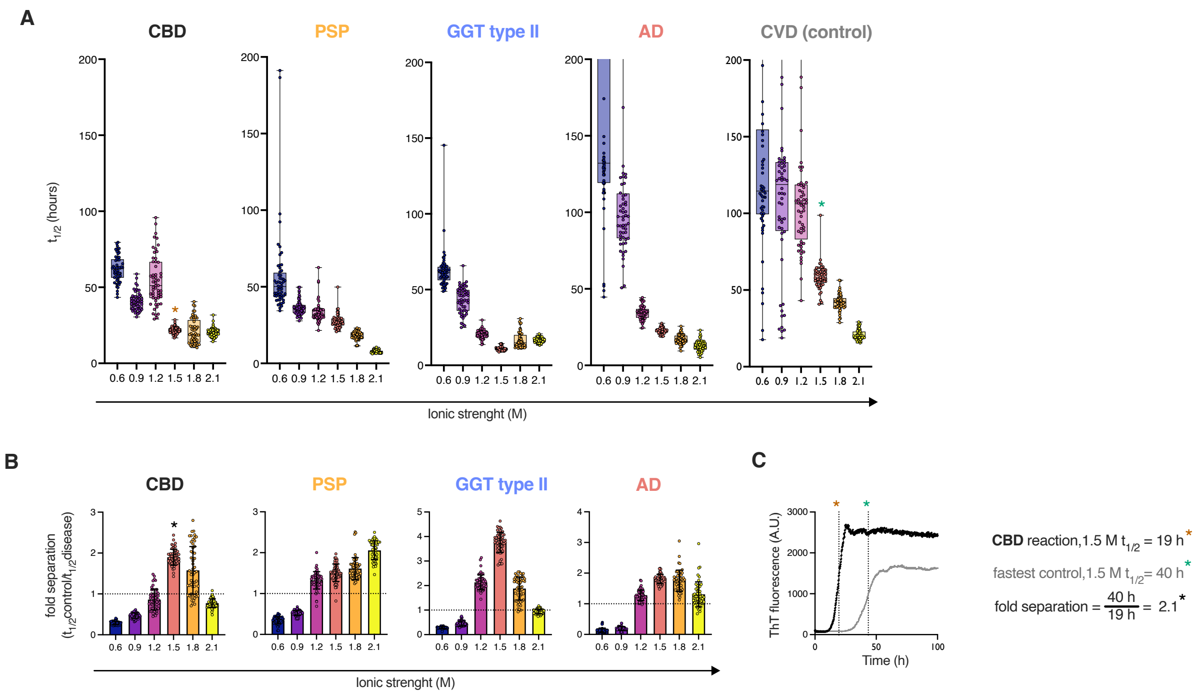


**Figure S3. Optimisation of K11 tau RT-QuIC: Ionic strength titration with sodium sulfate. (A)** Aggregation half-time ($t_{1/2}$) analysis of brain homogenate-seeded K11 reactions across a range of sodium sulfate concentrations, reported as ionic strength (M); boxes represent inter-quartile range, whiskers represent outer-quartiles. **(B)** Fold separation analysis, calculated as the ratio of the fastest $t_{1/2}$ control value divided by tauopathy-seeded $t_{1/2}$ values from (**A**); error bars represent standard deviation; the dashed line indicates the minimum fold separation (1) required to distinguish seeded from spontaneous aggregation. **(C)** Example fold separation calculation for a CBD-seeded reaction, illustrating the division of the fastest control $t_{1/2}$ (40 h) by a representative CBD $t_{1/2}$ at 1.5 M ionic strength. Asterisks in (**C**) mark the corresponding data points shown in (**A**) and (**B**). Brain samples used in this figure include 203, 209, MC24, AD2, and CVD1.

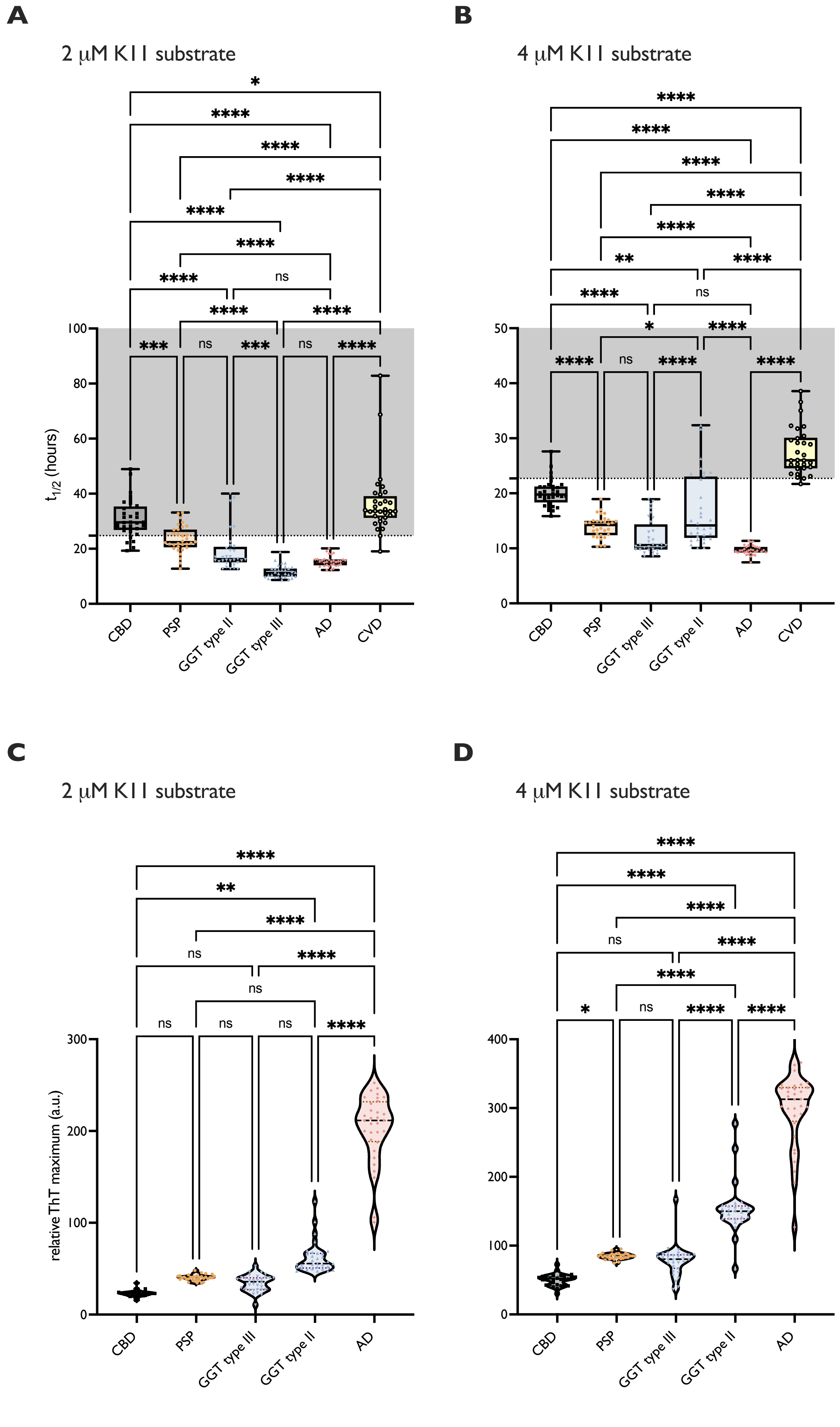
A
2 µM K11 substrate
t1/2 (hours)
CBD
PSP
GGT type II
GGT type III
AD
CVD
B
4 µM K11 substrate
C
2 µM K11 substrate
relative ThT maximum (a.u.)
D
4 µM K11 substrate

**Figure S4. Optimisation of K11 tau RT-QuIC: substrate concentration and strain discrimination**. (**A, B**) $t_{1/2}$ analysis of brain homogenate-seeded reactions compared to control CVD-seeded reactions in 2 μM K11 (A) and 4 μM K11 (B). (**C,D**) Relative ThT maxima from the reactions in (A) and (B). Asterisks represent statistical significance of one-way ANOVA with multiple comparisons; **** $p < 0.0001$, *** $p < 0.001$, ** $p < 0.01$, * $p < 0.1$, ns non-significant. Brain samples used in this figure include 203, 209, MC21, MC24, AD2, and CVD1.

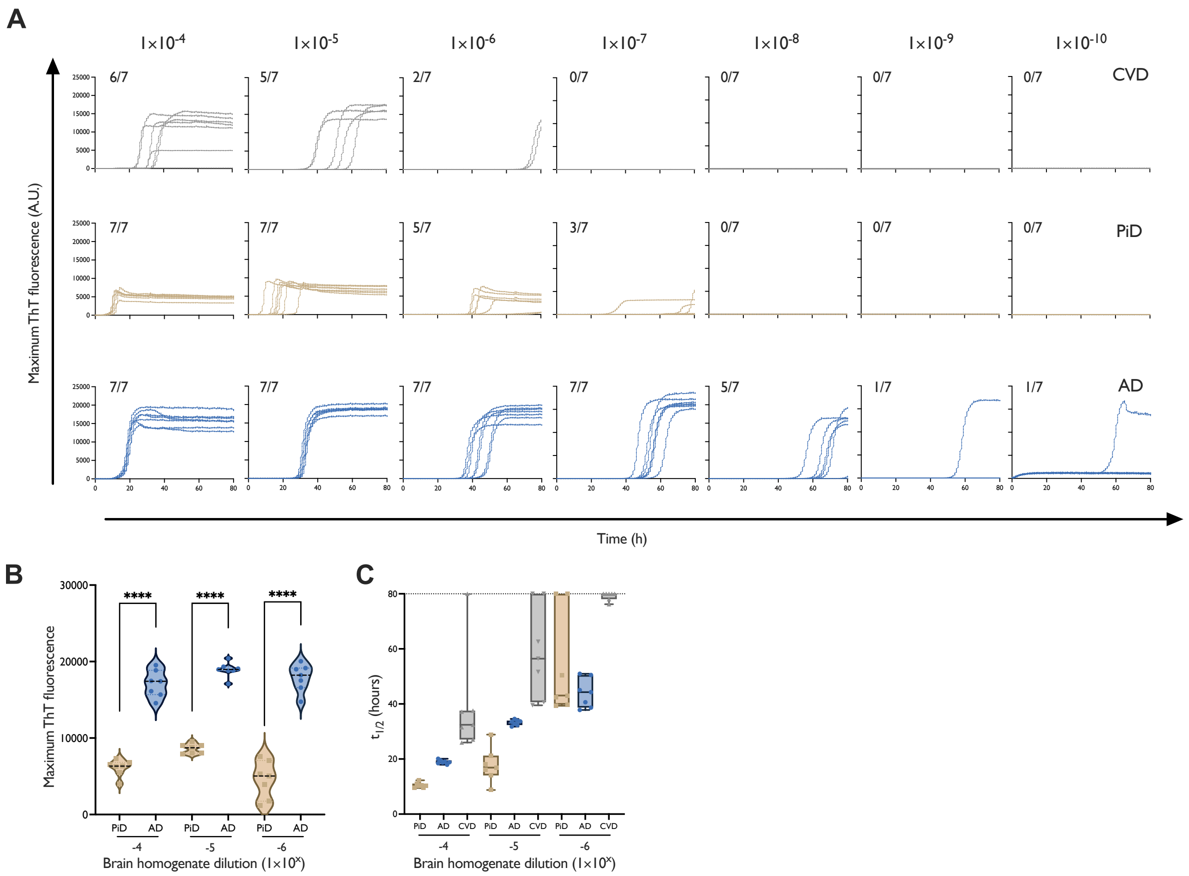


**Figure S5. Primary ThT fluorescence data of K12 RT-QuIC endpoint dilution analyses.** Panels show traces from 7 replicate reactions at the designated dilutions of cerebrovascular disease (CVD1), Alzheimer disease (AD6), and Pick disease (PiD6) brain homogenates in 250 mM $Na_3$citrate. The fractions in the upper left corner of each panel indicate the ThT-positive/total replicate reactions. (**B**) Violin plots of ThT maxima from reactions in (A) focusing on dilutions between $1x10^{-4}$ and $1x10^{-6}$. Statistical significance was assessed by one-way ANOVA; **** $p < 0.0001$; *** $p < 0.001$; ns, not significant. (**C**) box-and-whisker plots of $t_{1/2}$ values from reactions in (**A**); boxes represent inter-quartile range, whiskers represent outer quartiles.

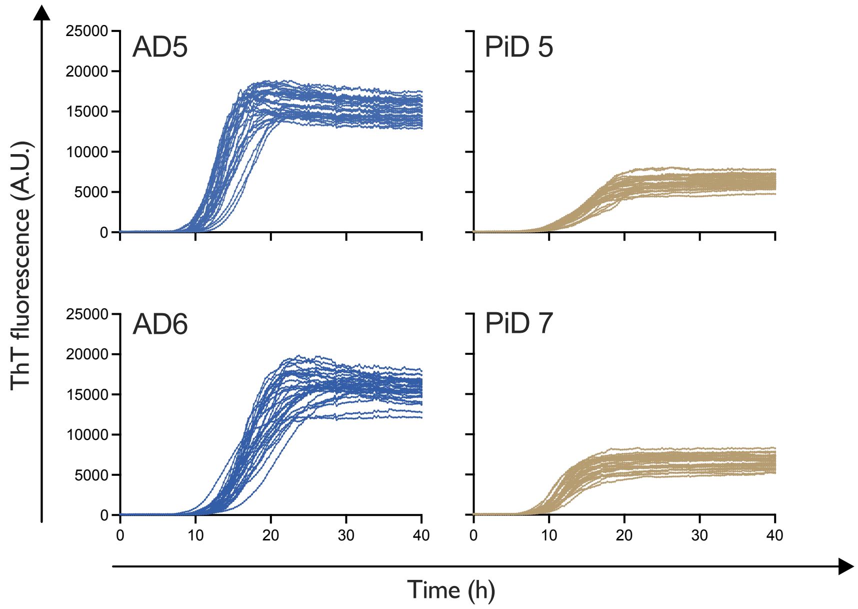


**Figure S6. K12 RT-QuIC raw fluorescence traces in 250 mM sodium citrate**. Individual panels represent a singular patient brain assayed at 1x10$^{-5}$ dilution, with 30 replicates per panel.

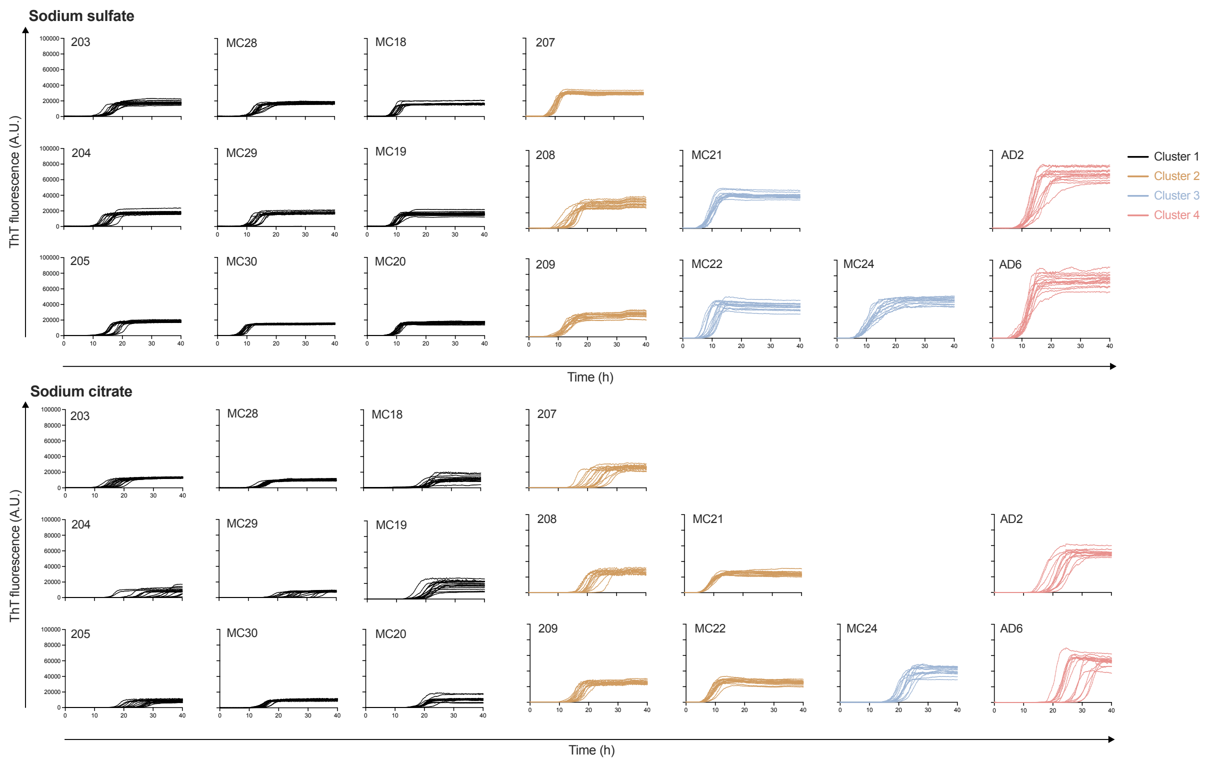


**Figure S7. K11 RT-QuIC raw fluorescence traces**. Traces are shown in 500 mM sodium sulfate (top) and 250 mM sodium citrate (bottom). Individual panels represent a singular patient brain assayed at $1x10^{-4}$ dilution, with sixteen replicates per panel.

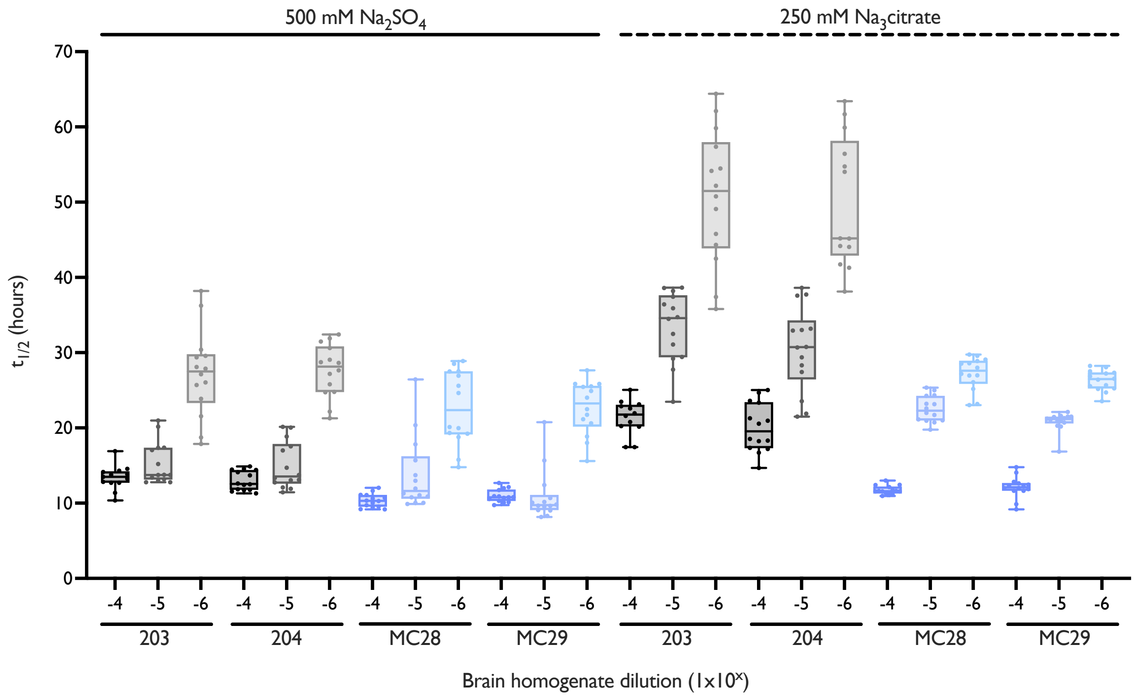


**Figure S8. Serial dilution kinetics of CBD and AGD-seeded K11 tau RT-QuIC reactions**. CBD (203,204) and AGD (MC28, M28) brain homogenates were assayed at 10-fold dilutions between $1x10^{-4}$ and $1x10^{-6}$ where each box represents a singular reaction condition, with sixteen replicate reactions per condition. The leftmost twelve reactions were performed in 500 mM sodium sulfate, rightmost twelve reactions in 250 mM sodium citrate; boxes represent inter-quartile range, whiskers represent outer quartiles.
